# Temporal Trends in Antimicrobial Resistance Among Clinically Important Pathogens in Iraq, 2021–2024: Implications for Empirical Antimicrobial Therapy

**DOI:** 10.64898/2026.09.30.26364389

**Authors:** Omeed Darweesh, Hemn Ahmed, Haidar Kadhim Al-Jawadi, Ramiar Kamal Kheder, Noor Najim Alwiswasi, Sarah Mohammed Hameed, Hemen Faik Mohammed, Amanj Kurdi

## Abstract

To characterise temporal changes in antimicrobial resistance (AMR) among clinically important bacterial pathogens in Iraq during 2021–2024 and examine their implications for empirical antimicrobial therapy and stewardship. We conducted a retrospective longitudinal analysis of aggregated national AMR surveillance data for *Escherichia coli, Klebsiella pneumoniae, Pseudomonas aeruginosa* and *Staphylococcus aureus*. Annual resistance proportions were evaluated across clinically relevant antimicrobials. Temporal change was characterised using absolute percentage-point changes and exploratory linear regression. Resistance trajectories were interpreted in relation to empirical treatment options and antimicrobial stewardship. The analysis included 92,338 organism records: 40,496 (43.9%) *E. coli*, 24,621 (26.7%) *S. aureus*, 18,223 (19.7%) *K. pneumoniae* and 8,998 (9.7%) *P. aeruginosa*. In 2024, ceftriaxone resistance reached 82.9% in *E. coli* and 65.2% in *K. pneumoniae*, while ciprofloxacin resistance was 55.4% and 57.9%, respectively. Meropenem resistance reached 37.4% in *K. pneumoniae* and 62.3% in *P. aeruginosa*. Conversely, resistance declined for amoxicillin–clavulanate in *E. coli* (83.5% in 2021 to 57.2% in 2024) and piperacillin–tazobactam in *P. aeruginosa* (46.4% to 32.9%). Several pathogen–antimicrobial combinations showed marked year-to-year variation, and no temporal trend remained statistically significant after false discovery rate correction. Resistance to several agents central to empirical therapy remains high in Iraq, while substantial carbapenem resistance constrains reliance on carbapenem escalation when resistance to other β-lactams is suspected. Heterogeneous trajectories indicate that empirical recommendations should not rely on single-year national antibiograms alone and support regular integration of national and local resistance surveillance into empirical treatment and antimicrobial stewardship strategies.

## Introduction

Antimicrobial resistance (AMR) increasingly threatens the effectiveness of empirical antibiotic therapy, particularly when treatment must be initiated before pathogen identification and antimicrobial susceptibility results are available. In 2019, bacterial AMR was estimated to be directly responsible for 1.27 million deaths and associated with 4.95 million deaths globally [1]. Preserving effective empirical treatment while minimising unnecessary exposure to broad-spectrum antibiotics therefore represents a key challenge for antimicrobial stewardship programmes. The World Health Organization (WHO) AWaRe Antibiotic Book provides guidance on antibiotic choice, dose, route and duration for more than 30 common infections [2, 3]. Its recommendations support preferential use of Access antibiotics where appropriate while limiting unnecessary exposure to Watch and Reserve agents. However, empirical antibiotic recommendations cannot be considered independently of local resistance epidemiology. Where resistance to recommended agents is high, unnecessary escalation to broader-spectrum Watch or Reserve antibiotics may also compromise stewardship. Longitudinal surveillance can therefore complement single-year antibiograms by showing whether resistance to clinically important antimicrobial agents is increasing, declining or fluctuating over time.

This issue is particularly relevant in Iraq, where recent surveillance has demonstrated high resistance among clinically important bacterial pathogens. A national analysis of 238,973 clinical specimens during 2020–2023 reported persistently high resistance to third-generation cephalosporins in *Escherichia coli*, substantial carbapenem resistance in *Klebsiella pneumoniae* and *Pseudomonas aeruginosa*, and considerable methicillin resistance in *Staphylococcus aureus* [4]. A multicentre study from four Iraqi governorates demonstrated substantial multidrug resistance [5]. Recent evidence also indicates challenges in antimicrobial use in Iraq [6–8]. Simulated-client studies reported non-prescription antibiotic dispensing in 80.6% of community-pharmacy encounters and inappropriate antibiotic supply in 60.0% of nurse-led clinic encounters, with substantial use of Watch antibiotics [7, 8]. These findings highlight concurrent resistance and antimicrobial-use challenges that may compromise empirical treatment and stewardship.

Although national Iraqi surveillance has described annual resistance patterns during 2020–2023 [4], longitudinal evaluation through 2024 is needed to determine whether resistance trajectories are consistent or heterogeneous and how they may constrain antimicrobial selection. We therefore analysed national AMR surveillance data from Iraq during 2021–2024 to characterise temporal changes among *E. coli, K. pneumoniae, P. aeruginosa* and *S. aureus* across clinically relevant antimicrobial agents. We aimed to identify resistance trajectories relevant to empirical antimicrobial therapy and to consider their implications for antimicrobial selection and stewardship, including WHO AWaRe recommendations.

## Materials and Methods

### Study Design and Data Source

We conducted a retrospective longitudinal analysis of national AMR surveillance data from Iraq during 2021–2024. Data were obtained from annual national antibiogram reports compiled by the Department of Clinical Pharmacy, Iraqi Ministry of Health and were accessed for research purposes on 11^th^ January 2026. The antibiograms collated antimicrobial susceptibility testing (AST) results from clinical microbiology laboratories across Iraqi governorates. The analysis used aggregated annual surveillance data rather than patient- or isolate-level records. The antibiograms reported annual organism counts and resistance percentages for individual antimicrobial agents. Antibiotic-specific numbers tested and resistant were unavailable; resistance percentages were therefore analysed as reported, while organism counts were used only to describe the surveillance population. Consequently, infection syndrome, specimen-specific resistance, healthcare setting, previous antimicrobial exposure and treatment outcomes were not available. Patient-level deduplication and co-resistance also could not be independently assessed. No formal sample-size calculation was performed because all available records for the four prespecified pathogens during 2021–2024 were included.

### Pathogens and antimicrobial agents

The analysis focused on four clinically important bacterial pathogens: *Escherichia coli*, *Klebsiella pneumoniae*, *Pseudomonas aeruginosa* and *Staphylococcus aureus*. These organisms were selected a priori because they represent major Gram-negative and Gram-positive causes of community- and healthcare-associated infections, are included within the WHO bacterial priority pathogens framework through clinically important resistant phenotypes, and had sufficiently consistent national surveillance data across 2021–2024 to permit longitudinal evaluation [9]. Their resistance profiles are also directly relevant to empirical antimicrobial selection for common and severe bacterial infections, including urinary tract infection, bloodstream infection, pneumonia and other healthcare-associated infections [2, 9]. Antimicrobial agents were grouped into clinically relevant classes according to the structure of the source surveillance data, including penicillins, cephalosporins, carbapenems, aminoglycosides, fluoroquinolones, macrolides, glycopeptides and tetracyclines, where applicable to the individual pathogen. Pathogen–antimicrobial combinations were included where resistance data were available for at least one surveillance year; analyses of temporal change used the years for which a resistance estimate was reported, whereas exploratory regression analyses were restricted to combinations with complete four-year data. Analyses were performed at the level of individual antimicrobial agents, with antimicrobial classes used to organise and interpret related resistance trajectories. Antibiotics were additionally considered in relation to their Access, Watch or Reserve classification and their role in empirical treatment recommendations in the WHO AWaRe Antibiotic Book [2, 3]. This classification was used for clinical and stewardship interpretation and did not alter the underlying resistance definitions.

### Microbiological testing and antimicrobial susceptibility

Bacterial identification and AST were undertaken by participating clinical microbiology laboratories using routine microbiological methods according to local laboratory procedures. The annual national antibiograms referenced the contemporaneous Clinical and Laboratory Standards Institute (CLSI) M100 standards [10–13]. Because the present study analysed aggregated national surveillance reports rather than original isolate-level laboratory records, susceptibility classifications and resistance percentages were analysed as reported by the surveillance system. The source datasets did not contain zone diameters, minimum inhibitory concentrations (MICs), or antibiotic-specific numbers tested and resistant. No retrospective reinterpretation of zone diameters or MICs, reclassification of susceptibility categories, or recalculation of resistance percentages was undertaken. Changes in CLSI interpretive criteria across surveillance years could not be retrospectively harmonised at isolate level.

### Statistical analysis

Resistance proportions were summarised for each pathogen–antimicrobial combination by calendar year as reported in the national antibiograms. Temporal change was characterised using the absolute change in resistance between 2021 and 2024, expressed in percentage points, together with the direction and consistency of annual changes. For combinations without resistance estimates in both 2021 and 2024, endpoint change was not calculated. Missing antimicrobial–year observations were retained as missing, with no imputation or interpolation. Exploratory linear regression was used to quantify temporal trends, with calendar year entered as the independent variable and annual resistance percentage as the dependent variable. Regression analyses were restricted to pathogen–antimicrobial combinations with resistance estimates available for all four years. All combinations meeting this completeness criterion were included in the regression analysis as reported in the source antibiograms; no post-hoc exclusions were made on the basis of the expected clinical applicability of individual pathogen–antimicrobial combinations. The regression coefficient (β) represented the estimated average annual change in resistance in percentage points per year. Positive coefficients indicated increasing resistance and negative coefficients indicated decreasing resistance. Because each regression was based on only four annual observations, trend estimates and associated statistical tests were interpreted cautiously. Emphasis was placed on the magnitude, direction and consistency of resistance change rather than statistical significance alone.

Given the number of pathogen–antimicrobial comparisons, Benjamini–Hochberg false discovery rate (FDR) correction was applied to nominal regression P values across all complete four-year regression models [14]. Statistical significance was defined as a two-sided P<0.05; however, FDR-adjusted results were interpreted alongside the magnitude and consistency of observed changes. No causal interpretation was assigned to observed temporal associations. Confidence intervals around resistance estimates and denominator-weighted temporal analyses were not calculated because antibiotic-specific numbers tested were unavailable in the source datasets. Analyses were conducted using R software.

### Interpretation in relation to empirical therapy and WHO AWaRe

To assess the potential clinical relevance of observed resistance trajectories, findings were interpreted according to the role of individual antibiotics in empirical treatment pathways, their spectrum of activity and their WHO AWaRe classification [2, 3]. Particular attention was given to resistance to agents commonly used for empirical treatment, changes affecting broad-spectrum and Watch antibiotics, and trajectories potentially relevant to carbapenem-sparing or other stewardship strategies. This analysis was intended to evaluate the microbiological reliability of potential empirical antimicrobial options at the population surveillance level. It did not assess individual treatment appropriateness, clinical response or treatment failure, and no resistance threshold was used to define an antimicrobial as clinically ineffective. Because the surveillance data were not stratified by infection syndrome, healthcare setting or individual patient characteristics, observed resistance percentages were not used to recommend specific empirical regimens. WHO AWaRe classification was used as an interpretive stewardship framework rather than as an outcome of the analysis.

### Ethical considerations

This study involved secondary analysis of de-identified, aggregated national antimicrobial resistance surveillance data provided by the Department of Clinical Pharmacy, Iraqi Ministry of Health, upon request. The dataset contained no patient-level or personally identifiable information, and the research team had no access to individual patient records. The authors did not have access to information that could identify individual participants at any stage during or after data collection. No additional specimens were collected and there was no direct involvement of patients or modification of clinical care. The analysis was conducted using existing aggregated surveillance data, and no individual informed consent was obtained because no identifiable or patient-level data were available to the research team. No separate research ethics approval was obtained for this secondary analysis. Reporting followed the Strengthening the Reporting of Observational Studies in Epidemiology (STROBE) statement, using the cross-sectional checklist as the closest applicable framework (**Appendix 1**) [15].

## Results

### Surveillance population

Across 2021–2024, the national antibiograms contained 92,338 organism records for the four study pathogens (Table 1), increasing from 3,215 records in 2021 to 34,884 in 2023 before decreasing to 30,038 in 2024. *Escherichia coli* accounted for 40,496 (43.9%) records, followed by *Staphylococcus aureus* (24,621; 26.7%), *Klebsiella pneumoniae* (18,223; 19.7%) and *Pseudomonas aeruginosa* (8,998; 9.7%).

**Table 1.** Annual surveillance population and distribution of the four study pathogens, Iraq, 2021–2024.

| Pathogen | 2021, n (%) | 2022, n (%) | 2023, n (%) | 2024, n (%) | 2021–2024, n (%) |
| --- | --- | --- | --- | --- | --- |
| *E. coli* | 1,525 (47.4) | 10,607 (43.8) | 16,218 (46.5) | 12,146 (40.4) | 40,496 (43.9) |
| *K. pneumoniae* | 600 (18.7) | 4,294 (17.7) | 7,393 (21.2) | 5,936 (19.8) | 18,223 (19.7) |
| *P. aeruginosa* | 343 (10.7) | 2,223 (9.2) | 2,752 (7.9) | 3,680 (12.3) | 8,998 (9.7) |
| *S. aureus* | 747 (23.2) | 7,077 (29.2) | 8,521 (24.4) | 8,276 (27.6) | 24,621 (26.7) |
| **Total** | 3,215 (100) | 24,201 (100) | 34,884 (100) | 30,038 (100) | 92,338 (100) |

(Note) Values are n (%). Percentages within each calendar year represent the proportion of the total organism records contributed by each pathogen. The 2021–2024 percentages represent each pathogen's proportion of the overall surveillance population. Percentages may not total exactly 100% because of rounding

### Temporal resistance patterns in *Escherichia coli*

Resistance to several β-lactams remained high (**Figure 1**, **Supplementary File 1**). Ceftriaxone resistance varied from 65.0% in 2021 to 83.4% in 2022, 68.9% in 2023 and 82.9% in 2024 (+17.9 percentage points), while cefotaxime increased from 44.6% to 66.9%, with no 2022 estimate. In contrast, cefepime resistance decreased from 61.2% to 34.7% (−26.5 percentage points), while ceftazidime was similar in 2021 and 2024 (64.4% and 63.0%). Carbapenem resistance remained lower than third-generation cephalosporin resistance. Imipenem increased from 22.3% to 26.6% and meropenem from 16.0% to 25.5%, both peaking in 2023 (37.9% and 36.3%, respectively), while ertapenem decreased from 12.7% to 10.2%. Ciprofloxacin resistance remained high (50.2% to 55.4%), while levofloxacin showed an irregular trajectory. Resistance decreased for amikacin (26.5% to 23.1%), gentamicin (34.1% to 31.2%) and tobramycin (40.0% to 29.1%). Amoxicillin–clavulanate showed a marked decline from 83.5% to 57.2% (−26.3 percentage points).

**Figure 1.**
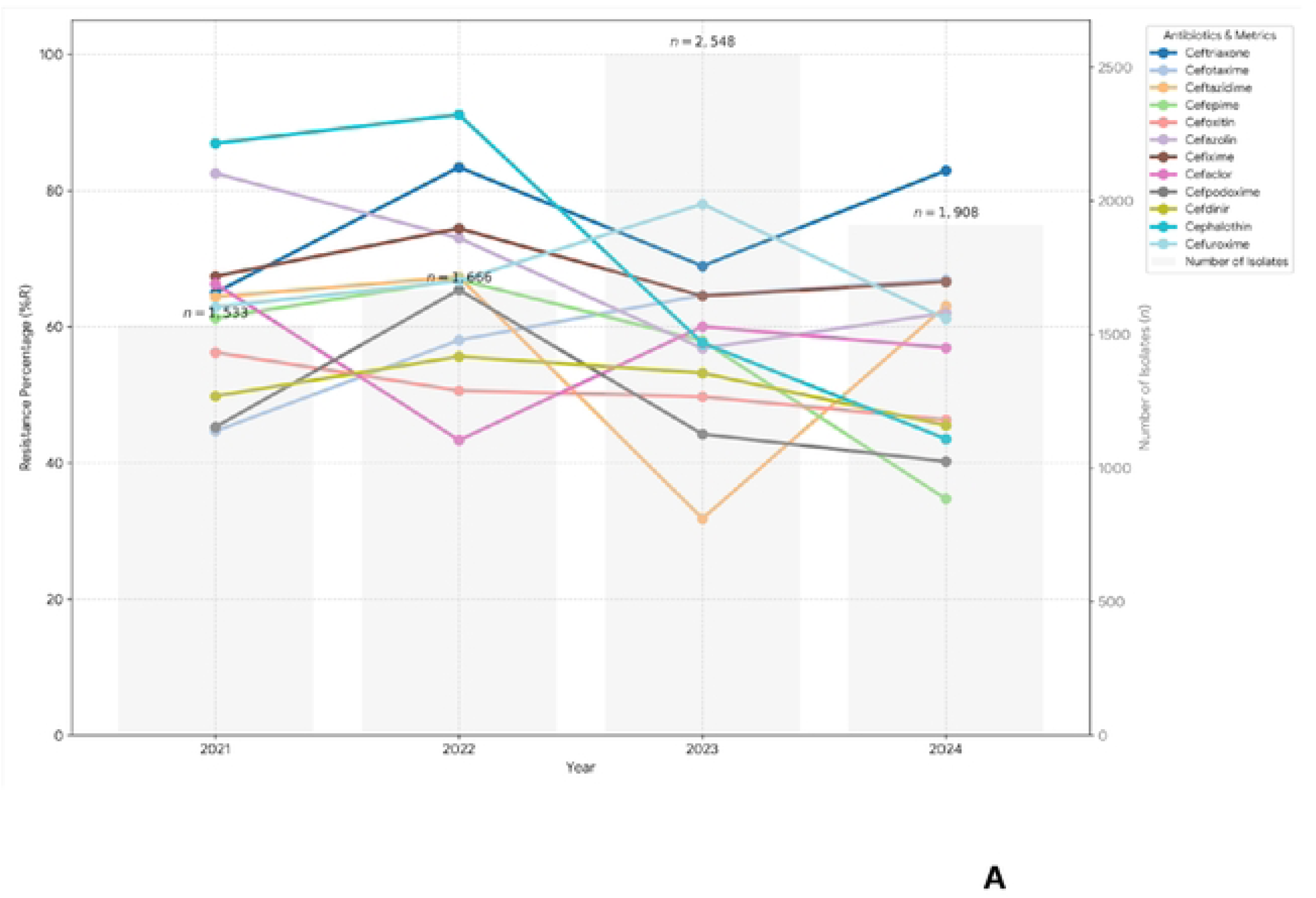
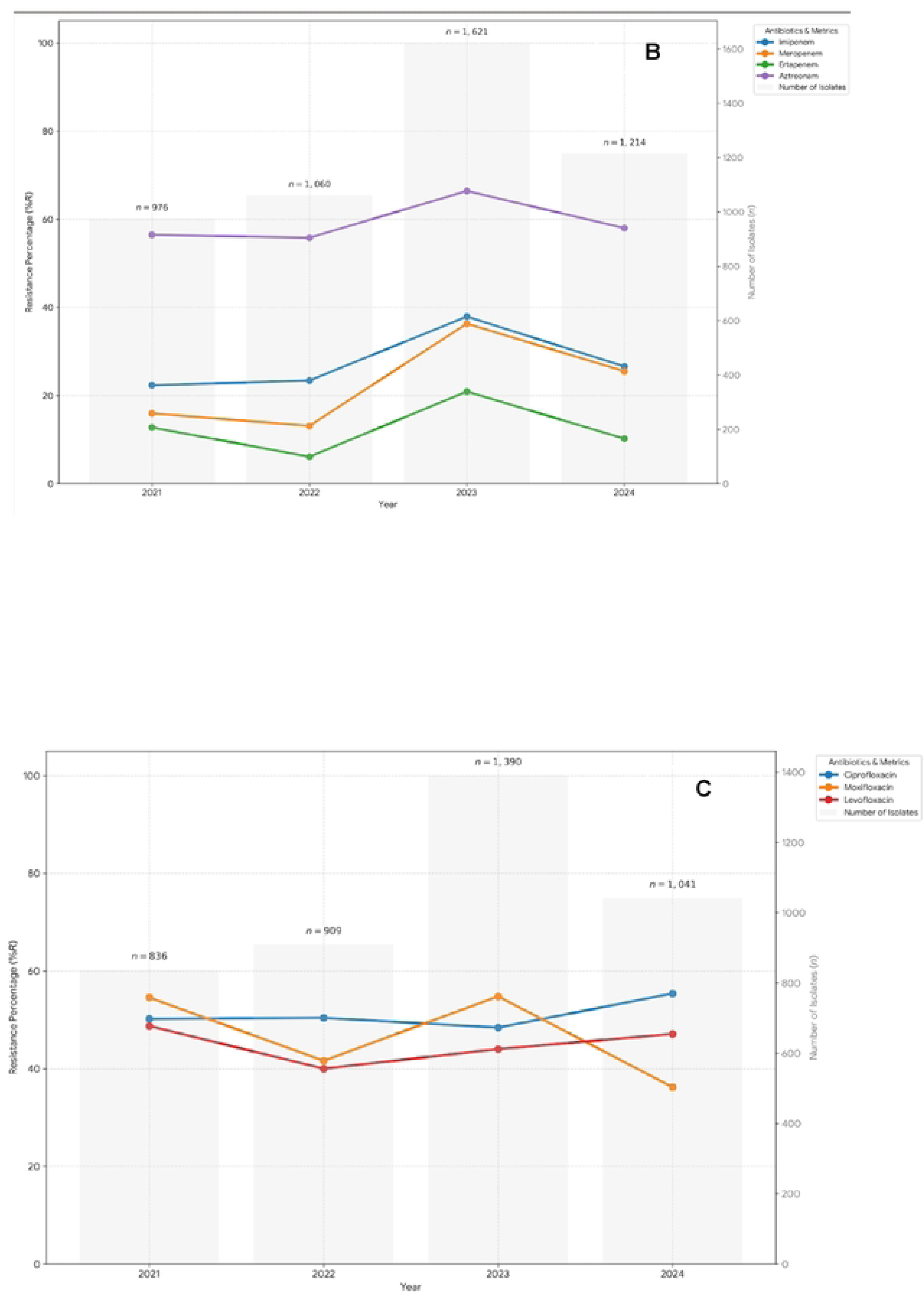
Resistance trajectories in *E. coli*, 2021–2024. Panels: **A, cephalosporins; B, carbapenems; C, fluoroquinolones** **General figure note:** Resistance percentages are those reported in the Iraqi annual national antibiograms for 2021-2024. Lines connect annual observations for visualisation of temporal patterns and should not be interpreted as continuous estimates between years. Any missing observations represent antimicrobial-year combinations for which resistance data were not reported in the source antibiogram. The figures are descriptive. Some organism-antimicrobial combinations included in the source surveillance data have limited clinical relevance or represent intrinsic/expected non-susceptibility and are presented for completeness rather than as potential therapeutic options.

### Temporal resistance patterns in *Klebsiella pneumoniae*

*K. pneumoniae* demonstrated sustained high resistance across several antimicrobial classes (**Figure 2**, **Supplementary File 2**). Ceftriaxone resistance increased from 51.6% in 2021 to 69.5% in 2022 and 90.0% in 2023, before decreasing to 65.2% in 2024, an overall increase of 13.6 percentage points between 2021 and 2024. Ceftazidime resistance decreased from 69.7% to 60.6%, while cefepime resistance declined from 80.6% to 58.4% (−22.2 percentage points). Carbapenem resistance remained substantial. Imipenem resistance was 46.4% in 2021 and 44.0% in 2024, while meropenem resistance decreased from 46.4% to 37.4%. In contrast, ertapenem resistance increased from 15.7% to 28.0%, representing an increase of 12.3 percentage points. Fluoroquinolone resistance increased overall. Ciprofloxacin resistance increased each year from 42.5% in 2021 to 50.5% in 2022, 56.8% in 2023 and 57.9% in 2024, an absolute increase of 15.4 percentage points. Levofloxacin resistance increased overall from 43.4% to 55.4%, although the annual trajectory was less consistent. Among aminoglycosides, amikacin resistance was 38.2% in 2021 and 42.7% in 2024, while gentamicin resistance increased from 49.6% to 55.6%. Piperacillin–tazobactam resistance decreased from 59.6% to 51.9% but remained above 50% in 2024.

**Figure 2.**
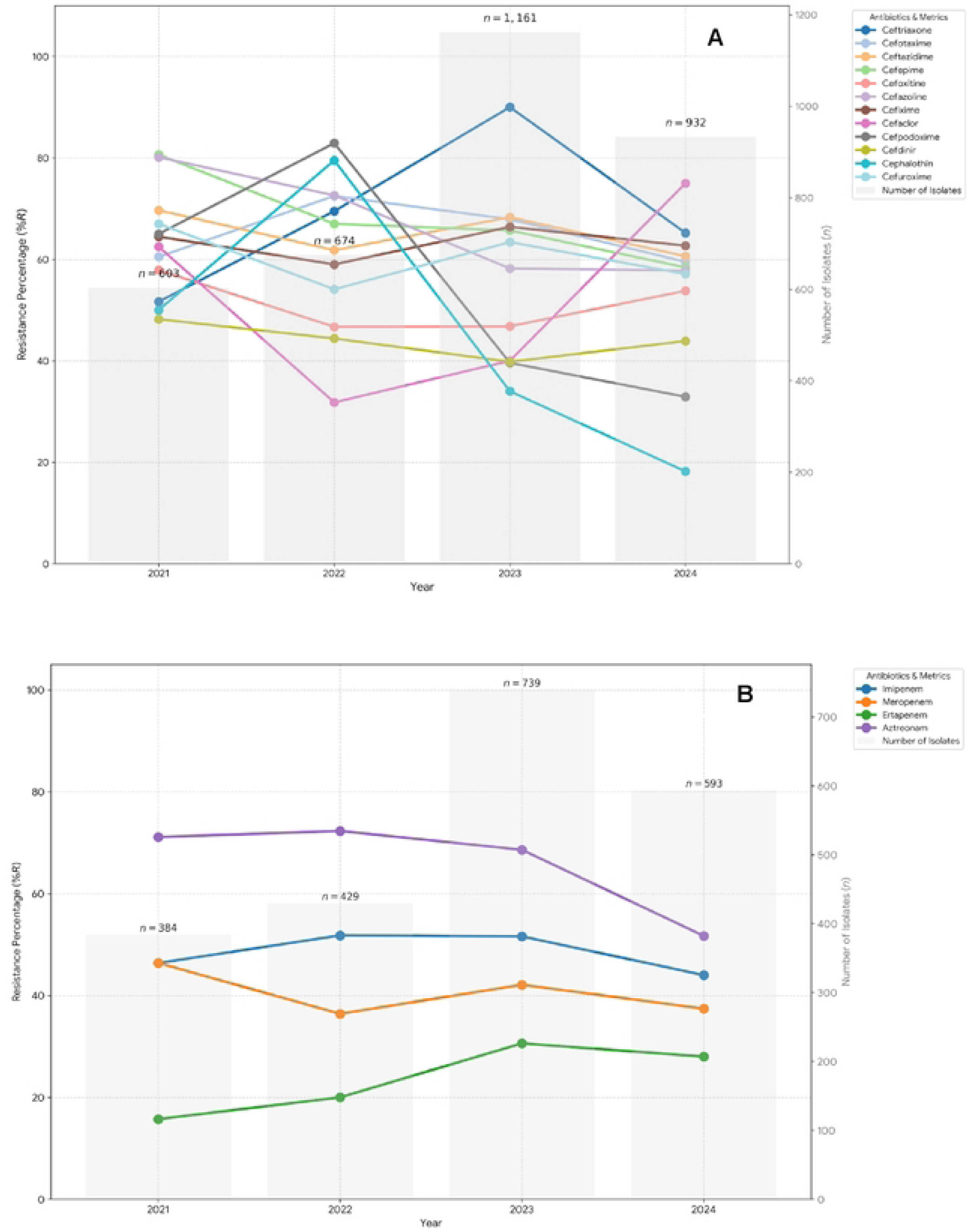
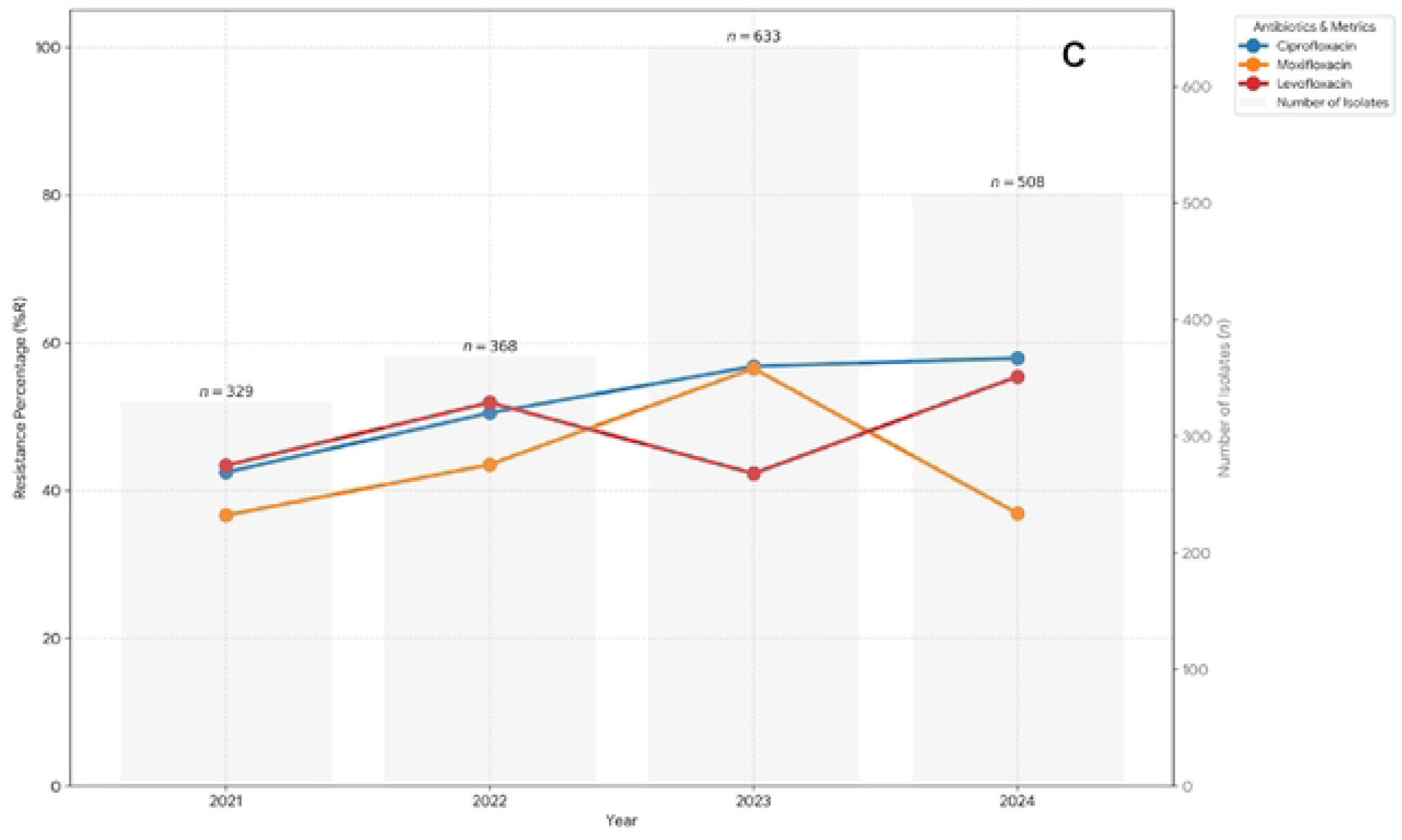
Resistance trajectories in *K. pneumoniae*, 2021–2024. Panels: A, cephalosporins; B, carbapenems; C, fluoroquinolones **General figure note:** Resistance percentages are those reported in the Iraqi annual national antibiograms for 2021-2024. Lines connect annual observations for visualisation of temporal patterns and should not be interpreted as continuous estimates between years. Any missing observations represent antimicrobial-year combinations for which resistance data were not reported in the source antibiogram. The figures are descriptive. Some organism-antimicrobial combinations included in the source surveillance data have limited clinical relevance or represent intrinsic/expected non-susceptibility and are presented for completeness rather than as potential therapeutic options.

### Temporal resistance patterns in *Pseudomonas aeruginosa*

Resistance among *P. aeruginosa* remained high across several major antipseudomonal antimicrobial classes (**Figure 3**, **Supplementary File 3**). Imipenem resistance increased from 49.8% in 2021 to 55.0% in 2024. Meropenem showed greater temporal variation, decreasing from 53.1% in 2021 to 45.9% in 2022 and 41.9% in 2023, before increasing to 62.3% in 2024, an overall increase of 9.2 percentage points. Ceftazidime resistance decreased from 63.9% in 2021 to 52.3% in 2024, while cefepime resistance decreased from 63.0% to 57.2%. Piperacillin–tazobactam showed a more consistent downward trajectory, with resistance decreasing from 46.4% in 2021 to 37.4% in 2022, 37.8% in 2023 and 32.9% in 2024, an absolute reduction of 13.5 percentage points. Fluoroquinolone resistance remained high and fluctuated considerably. Ciprofloxacin resistance decreased from 57.8% in 2021 to 39.2% in 2023 before increasing to 53.4% in 2024. Levofloxacin showed a similar pattern, decreasing from 58.9% in 2021 to 43.0% in 2023 before returning to 58.6% in 2024. Among aminoglycosides, amikacin resistance increased from 39.2% to 45.0%, gentamicin resistance decreased from 64.0% to 53.1%, and tobramycin resistance remained largely unchanged overall (57.8% versus 57.0%).

**Figure 3.**
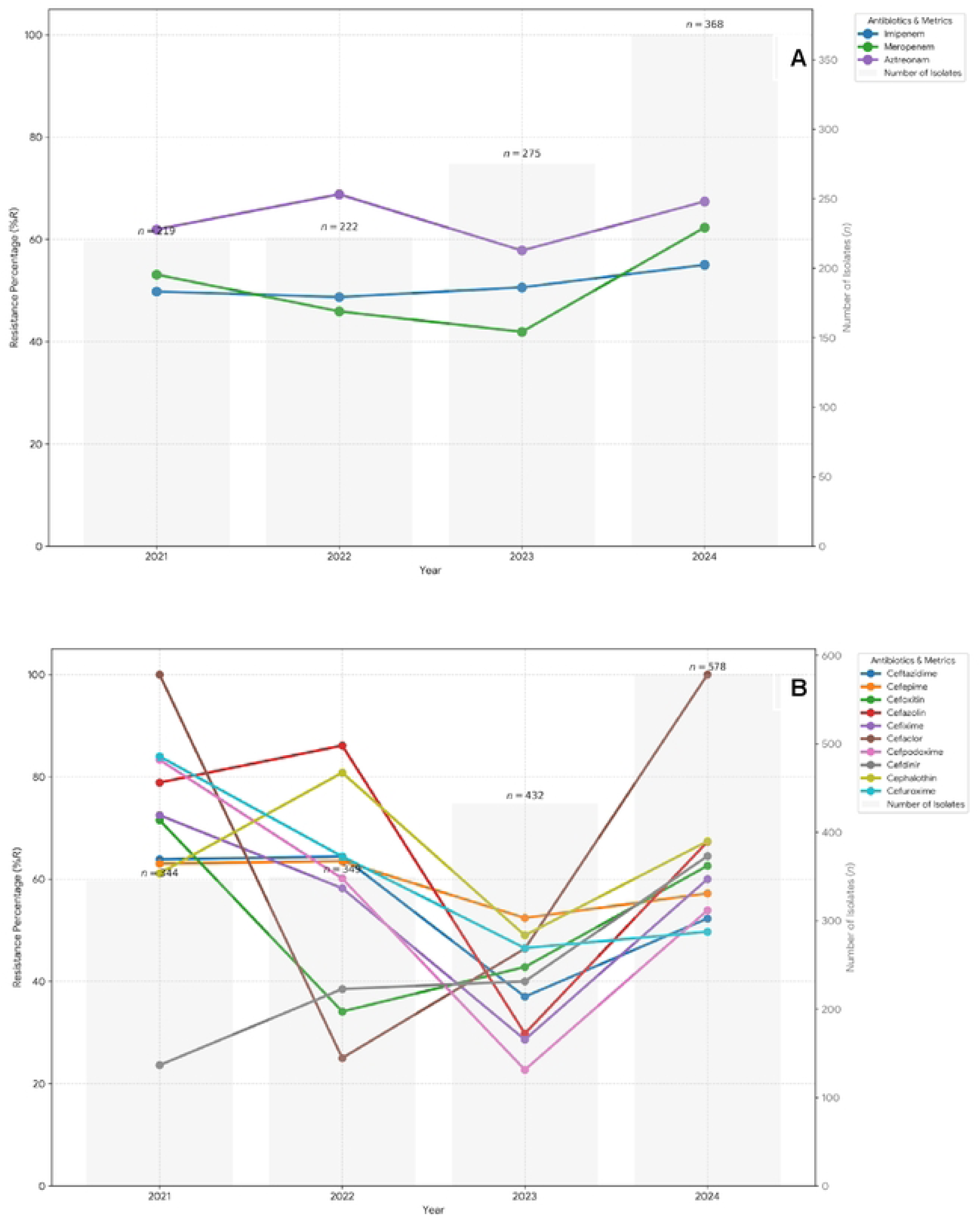
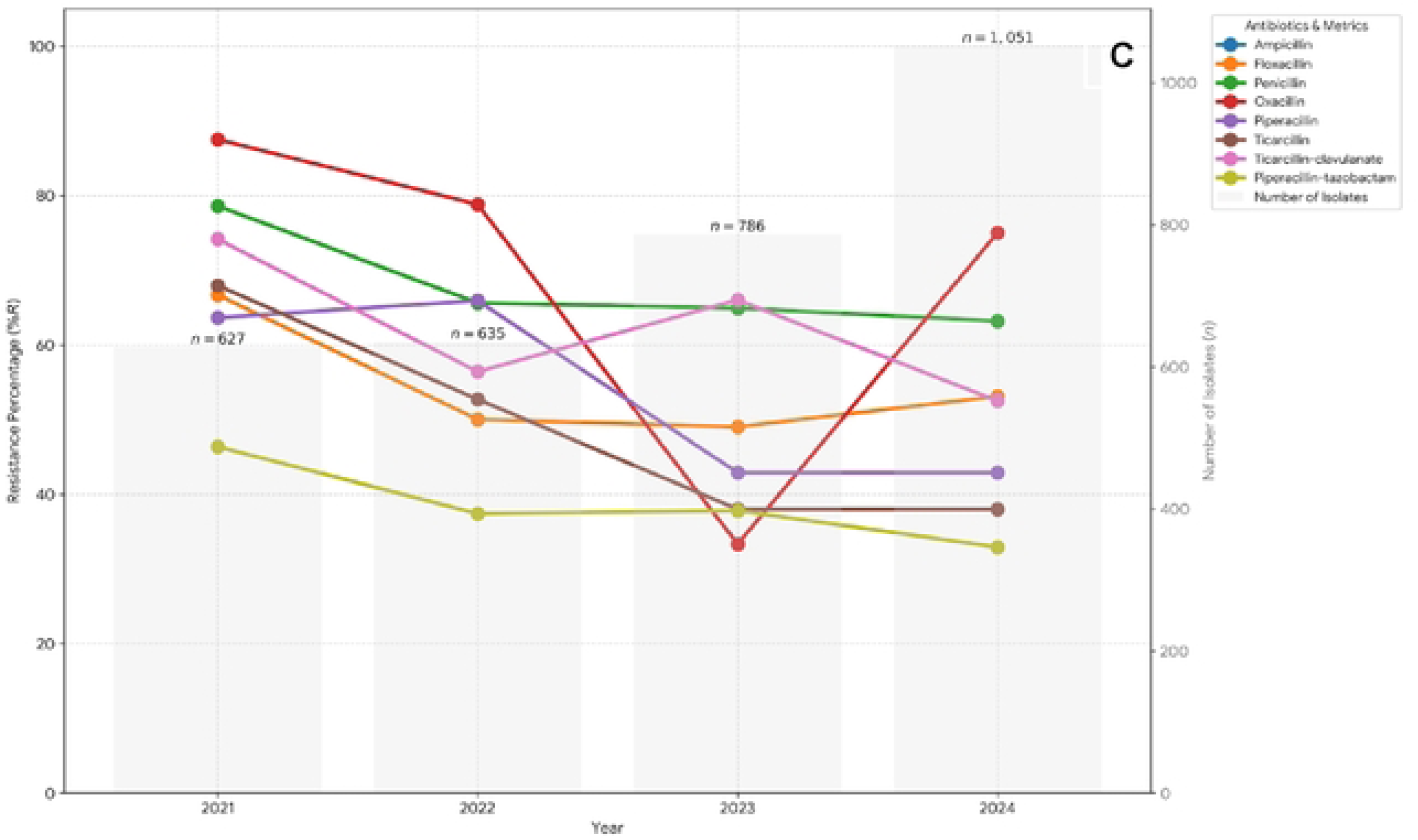
Resistance trajectories in *P. aeruginosa*, 2021–2024. Panels: **A, carbapenems; B, cephalosporins; C, penicillins** **General figure note:** Resistance percentages are those reported in the Iraqi annual national antibiograms for 2021-2024. Lines connect annual observations for visualisation of temporal patterns and should not be interpreted as continuous estimates between years. Any missing observations represent antimicrobial-year combinations for which resistance data were not reported in the source antibiogram. The figures are descriptive. Some organism-antimicrobial combinations included in the source surveillance data have limited clinical relevance or represent intrinsic/expected non-susceptibility and are presented for completeness rather than as potential therapeutic options.

### Temporal resistance patterns in *Staphylococcus aureus*

Resistance to penicillin remained consistently high in S. aureus, at 76.7% in 2021 and 78.3% in 2024 (**Figure 4**, **Supplementary File 4**). Oxacillin resistance was 78.5% in 2021, 65.1% in 2023 and 75.4% in 2024; no corresponding value was reported in 2022, precluding assessment of a complete four-year trajectory. Erythromycin resistance was also high, increasing from 61.2% in 2021 to 68.8% in 2022 and 69.5% in 2023, before decreasing to 66.5% in 2024. In contrast, several aminoglycosides showed substantial reductions in resistance. Amikacin resistance decreased progressively from 52.6% in 2021 to 31.8% in 2022, 26.9% in 2023 and 17.8% in 2024, corresponding to an absolute reduction of 34.8 percentage points. Tobramycin resistance decreased from 43.2% to 27.2%, while gentamicin resistance decreased overall from 41.3% to 31.5%, despite an increase to 61.4% in 2022. Ciprofloxacin resistance also decreased, from 51.5% in 2021 to 34.6% in 2024 (−16.9 percentage points), whereas levofloxacin resistance increased from 34.7% to 42.9%.

**Figure 4.**
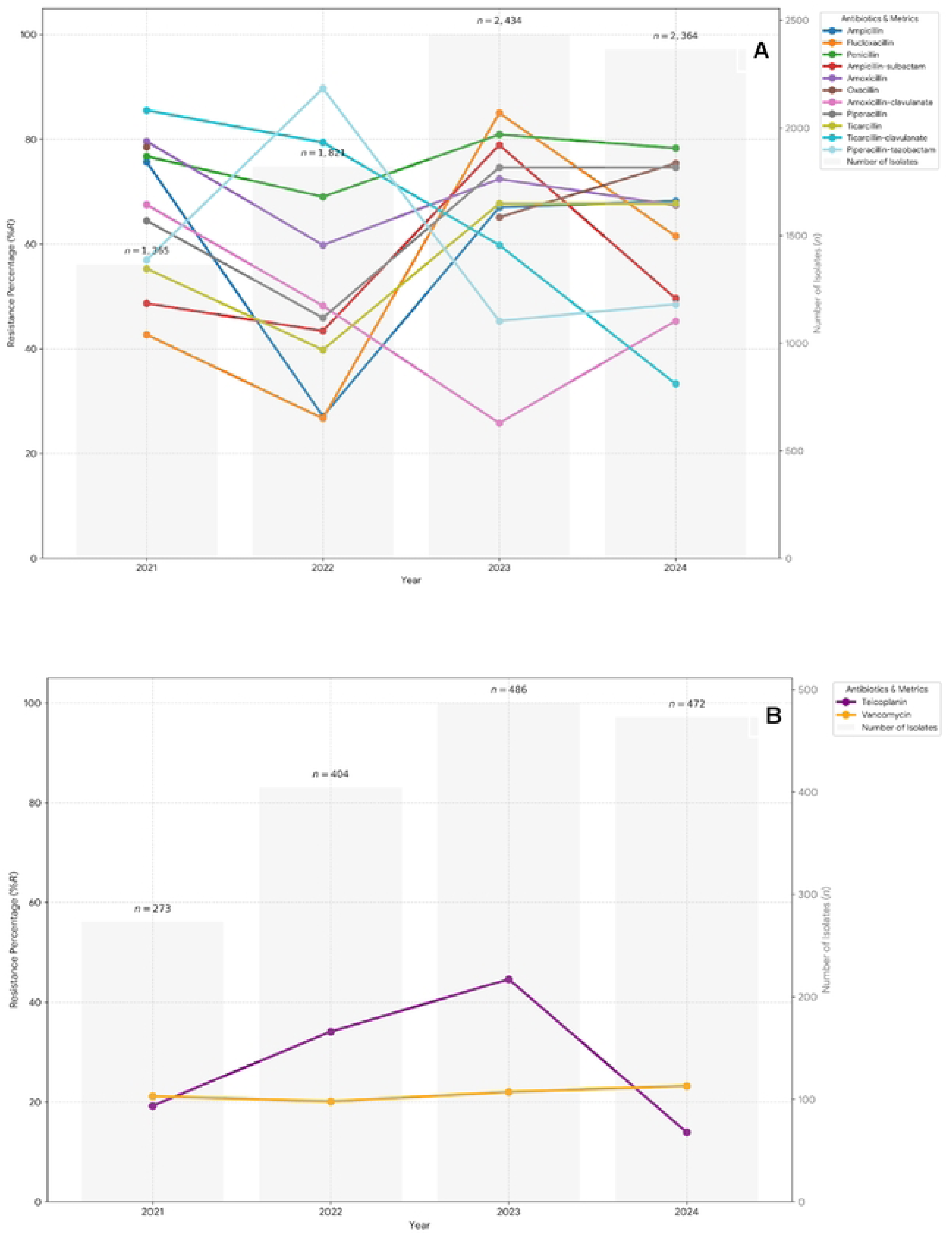
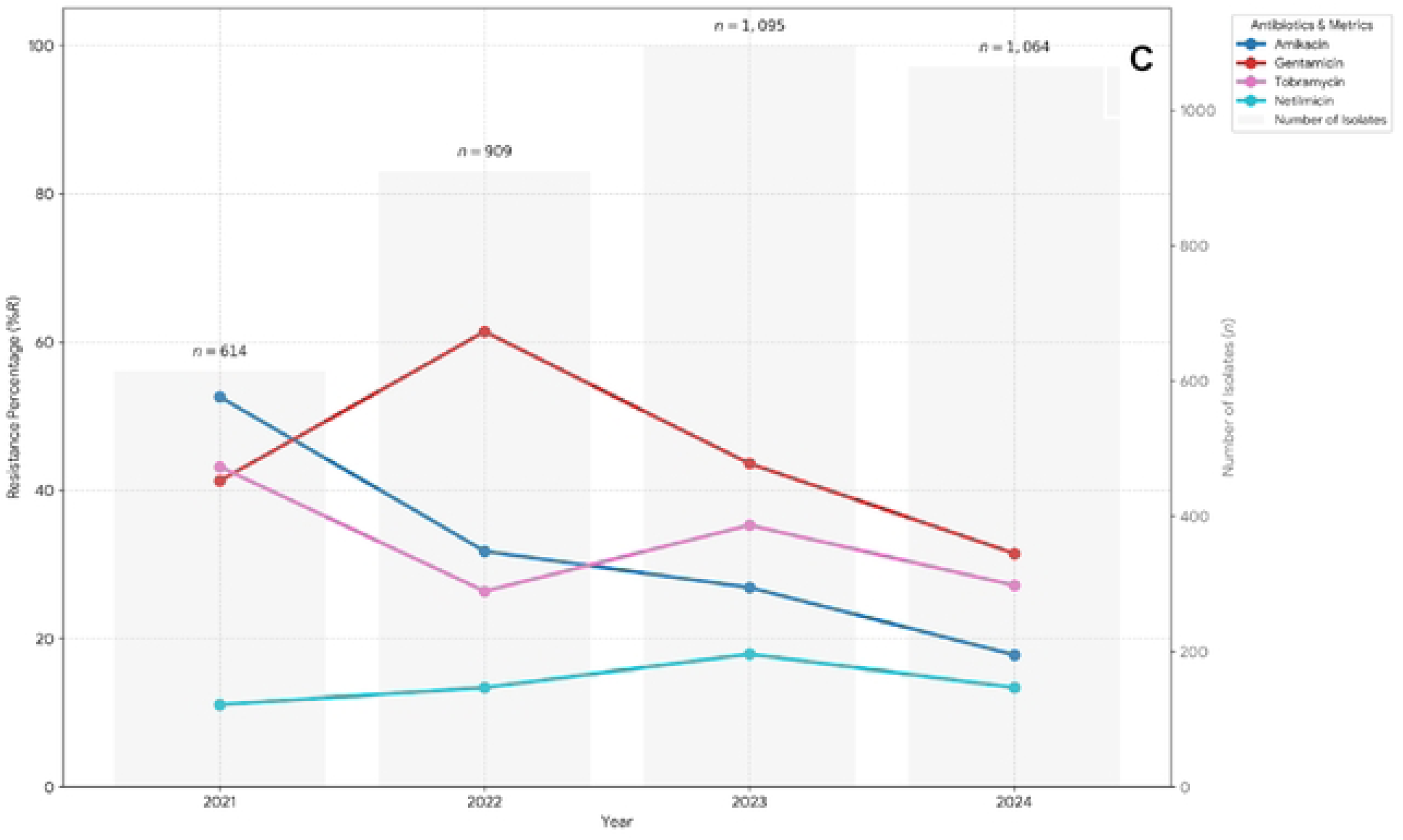
Resistance trajectories in *S. aureus*, 2021–2024. Panels**: A, penicillins/β-lactams; B, glycopeptides; C, aminoglycosides** **General figure note:** Resistance percentages are those reported in the Iraqi annual national antibiograms for 2021-2024. Lines connect annual observations for visualisation of temporal patterns and should not be interpreted as continuous estimates between years. Any missing observations represent antimicrobial-year combinations for which resistance data were not reported in the source antibiogram. The figures are descriptive. Some organism-antimicrobial combinations included in the source surveillance data have limited clinical relevance or represent intrinsic/expected non-susceptibility and are presented for completeness rather than as potential therapeutic options.

Vancomycin resistance was reported as 21.2% in 2021, 20.1% in 2022 and 23.2% in 2024; no estimate was reported for 2023. These values are presented as reported in the national antibiograms. In the absence of isolate-level MIC or confirmatory susceptibility data, they were treated as an unconfirmed surveillance signal rather than as estimates of the prevalence of confirmed vancomycin-resistant *S. aureus*. Tetracycline resistance increased from 47.8% in 2021 to 68.3% in 2024 (+20.5 percentage points), while doxycycline resistance changed from 46.8% to 40.6%, with lower values reported during the intervening years.

### Exploratory temporal trend analysis

Temporal resistance trajectories were heterogeneous, with several pathogen–antimicrobial combinations demonstrating substantial year-to-year variation. Exploratory linear regression was restricted to combinations with resistance estimates available for all four years. Of the 181 complete pathogen–antimicrobial series evaluated, 12 (6.6%) yielded nominal P values <0.05, but none remained statistically significant after Benjamini–Hochberg false discovery rate correction. The 181 models comprised all pathogen–antimicrobial combinations meeting the four-year completeness criterion in the source antibiograms, irrespective of their clinical applicability. Given that each model contained only four annual observations, these analyses were considered exploratory, and greater emphasis was placed on the magnitude, direction and consistency of resistance change. The clearest directional patterns among clinically relevant combinations included increasing ciprofloxacin resistance in *K. pneumoniae* (42.5% to 57.9%; +15.4 percentage points), decreasing piperacillin–tazobactam resistance in *P. aeruginosa* (46.4% to 32.9%; −13.5 percentage points), decreasing amoxicillin– clavulanate resistance in *E. coli* (83.5% to 57.2%; −26.3 percentage points), and progressively decreasing amikacin resistance in *S. aureus* (52.6% to 17.8%; −34.8 percentage points). However, marked annual fluctuations in several cephalosporin, carbapenem and fluoroquinolone series indicated that endpoint changes did not invariably represent consistent linear trends.

### Discussion

#### Principal findings

In this national longitudinal analysis of 92,338 organism records from Iraq between 2021 and 2024, resistance remained high across several antimicrobial classes central to empirical treatment, but trajectories differed substantially by pathogen and antimicrobial agent. High third-generation cephalosporin and fluoroquinolone resistance in *E. coli* and *K. pneumoniae* constrained commonly used empirical options, while substantial carbapenem resistance in *K. pneumoniae* and *P. aeruginosa* indicated that escalation to carbapenems cannot be assumed to overcome resistance. Conversely, declining resistance was observed for amoxicillin–clavulanate in *E. coli*, piperacillin–tazobactam in *P. aeruginosa*, and several aminoglycosides in *S. aureus*. However, marked year-to-year variation cautions against interpreting endpoint changes as stable resistance trends.

These findings extend recent Iraqi surveillance reporting of high third-generation cephalosporin and carbapenem resistance by incorporating 2024 data and evaluating longitudinal trajectories [4]. None of the temporal trends remained statistically significant after correction for multiple comparisons, supporting interpretation based primarily on their magnitude, direction and consistency rather than statistical significance alone.

#### Implications for empirical treatment of Enterobacterales

The resistance profiles of *E. coli* and *K. pneumoniae* present challenges for empirical treatment. Ceftriaxone resistance was 82.9% in *E. coli* and 65.2% in *K. pneumoniae* in 2024, while ciprofloxacin resistance was 55.4% and 57.9%, respectively. These levels raise concern about empirical reliance on these agents, although the absence of syndrome- and setting-specific data precludes estimation of empirical coverage. Third-generation cephalosporin-resistant and carbapenem-resistant Enterobacterales are classified as critical-priority pathogens in the WHO 2024 Bacterial Priority Pathogens List [9]. High cephalosporin resistance may encourage empirical carbapenem escalation, but carbapenem resistance was itself substantial. In *K. pneumoniae*, 2024 resistance was 44.0% to imipenem and 37.4% to meropenem, while meropenem resistance in *E. coli* was 25.5%. Amikacin resistance in *E. coli* was 23.1% in 2024, lower than ceftriaxone and ciprofloxacin. This does not support direct therapeutic substitution because antimicrobial selection depends on infection syndrome, patient factors, toxicity and local susceptibility. Rather, it illustrates that broader spectrum does not necessarily equate to more reliable microbiological activity.

#### Limited empirical options for *Pseudomonas aeruginosa*

The findings for *P. aeruginosa* indicate substantial constraints on conventional antipseudomonal options. In 2024, resistance exceeded 50% for meropenem, imipenem, ceftazidime, cefepime, ciprofloxacin, levofloxacin, gentamicin and tobramycin. Carbapenem-resistant *P. aeruginosa* is a high-priority pathogen in the 2024 WHO Bacterial Priority Pathogens List [9]. Piperacillin–tazobactam resistance declined from 46.4% to 32.9%, one of the more consistent downward trajectories, although the 2024 value does not establish reliable empirical activity. Marked variation in meropenem and fluoroquinolone resistance illustrates the limitations of basing empirical guidelines on a single annual antibiogram. Where *P. aeruginosa* is suspected, empirical selection should therefore incorporate current local susceptibility data and timely microbiological results where available.

#### *Staphylococcus aureus*: divergent trajectories and a vancomycin surveillance signal

Oxacillin resistance in *S. aureus* remained high where reported, reaching 75.4% in 2024. Methicillin-resistant *S. aureus* is classified as a high-priority pathogen by WHO [9]. Amikacin resistance declined progressively from 52.6% in 2021 to 17.8% in 2024, among the largest changes observed, whereas tetracycline resistance increased from 47.8% to 68.3%, illustrating divergent within-pathogen trajectories. The decline in amikacin resistance represents a microbiological trajectory and should not be interpreted as evidence supporting treatment substitution. Particular caution is required for vancomycin resistance, reported at approximately 20–23%. In the absence of isolate-level MIC and confirmatory testing, these values should be regarded as an unconfirmed surveillance signal requiring validation, rather than a prevalence estimate of confirmed vancomycin-resistant *S. aureus*. Vancomycin resistance has also been reported in a previous Iraqi surveillance study [5]. Confirmation using isolate-level MIC and appropriate confirmatory testing is therefore required

#### Implications for the Iraqi National Antimicrobial Guidelines and antimicrobial stewardship

The present findings have direct relevance to the Iraqi National Antimicrobial Guidelines (INAG) [16]. The observed resistance trajectories provide contemporary surveillance evidence relevant to future review of empirical recommendations. Ceftriaxone is included among INAG empirical options for several infections in which Enterobacterales may be implicated [16], whereas 2024 resistance was 82.9% in *E. coli* and 65.2% in *K. pneumoniae*. Carbapenems and antipseudomonal β-lactams also feature in selected empirical pathways, while substantial resistance to these agents was observed in *K. pneumoniae* and *P. aeruginosa*. These comparisons do not establish that individual INAG recommendations are inappropriate, because our data lack infection syndrome, specimen, healthcare setting and patient risk factors. Rather, they identify empirical options for which contemporary local and national susceptibility data are particularly important. High resistance to one empirical option should not automatically lead to a broader-spectrum agent. Substantial carbapenem resistance, particularly in *K. pneumoniae* and *P. aeruginosa*, supports combining national surveillance with local antibiograms, microbiological sampling and timely review of empirical therapy [16]. Similar considerations apply to the WHO AWaRe framework. AWaRe promotes preferential use of Access antibiotics where clinically appropriate [2, 3], but empirical recommendations require adaptation to local resistance epidemiology. In Iraq, stewardship must balance adequate empirical coverage against unnecessary use of broader-spectrum Watch and Reserve agents. AWaRe classification and local resistance surveillance should therefore be viewed as complementary. Recent simulated-client studies in Iraq identified frequent non-prescription or inappropriate antibiotic supply in community pharmacies and nurse-led clinics, including substantial use of Watch antibiotics [7, 8]. Stewardship should therefore extend beyond hospitals and integrate resistance surveillance with interventions addressing antimicrobial access, dispensing and use. Periodic review of INAG recommendations against contemporary national and local resistance data could therefore strengthen the translation of surveillance into empirical treatment guidance.

#### Implications for surveillance in Iraq

Beyond individual antimicrobial choices, the findings have implications for AMR surveillance in Iraq. National antibiograms provide valuable longitudinal information but would be strengthened by facility-level antibiograms and, where possible, stratification by specimen type, clinical setting and patient characteristics. More standardised surveillance, with consistent reporting of pathogen–antimicrobial testing denominators, would improve temporal interpretation and statistical precision. Integration of microbiology, prescribing and pharmacy data could enable assessment of relationships among resistance, antimicrobial use and clinical outcomes. Systematic linkage of surveillance with guideline development and stewardship could further strengthen translation into clinical practice.

#### Strengths and limitations

This study has several strengths. It analyses four consecutive years of national AMR surveillance data, includes more than 92,000 organism records and evaluates resistance trajectories across four clinically important bacterial pathogens and multiple antimicrobial classes. Its longitudinal design enables assessment of both persistently high resistance and temporal change beyond that provided by single-year antibiograms.

Several limitations should be acknowledged. First, the data lacked individual-level information on infection syndrome, specimen type, healthcare setting, prior antimicrobial exposure and clinical outcomes. Community- and healthcare-associated infection and colonisation could not be distinguished. Second, antibiotic-specific testing denominators were unavailable; consequently, confidence intervals and denominator-weighted analyses could not be calculated, and temporal analyses relied on reported annual percentages. Third, annual surveillance volume varied substantially, from 3,215 records in 2021 to 34,884 in 2023. Changes in surveillance coverage, participating laboratories, sampling or represented populations may therefore have contributed to apparent temporal changes and cannot be distinguished from changes in underlying resistance epidemiology. Fourth, patient-level deduplication could not be independently assessed; organism records therefore should not be interpreted as unique patients. Co-resistance and multidrug resistance also could not be assessed. Fifth, laboratory methods, antimicrobial panels and interpretive criteria may have varied across sites and over time. Although the antibiograms referenced contemporaneous CLSI standards [10–13]; isolate-level zone diameters and MIC data were unavailable, preventing retrospective harmonisation. Sixth, resistance data were not available for every antimicrobial in every year, reducing complete four-year series available for regression. Seventh, each regression was based on only four annual observations, limiting statistical power and sensitivity to non-linear trajectories. None of the observed trends remained statistically significant after FDR correction, and the trajectories should be considered descriptive and hypothesis-generating. Finally, resistance percentages cannot be translated directly into treatment recommendations or failure probabilities. They require integration with syndrome-specific evidence, local antibiograms and clinical judgement.

#### Conclusions

National AMR surveillance in Iraq during 2021–2024 demonstrated persistently high resistance to several antimicrobial agents relevant to empirical therapy, including third-generation cephalosporins and fluoroquinolones in *E. coli* and *K. pneumoniae*, together with substantial carbapenem resistance in *K. pneumoniae* and *P. aeruginosa*. These findings indicate that escalation towards broader-spectrum agents cannot be assumed to overcome resistance when commonly used empirical options are compromised. Resistance trajectories were heterogeneous, with both increasing and decreasing patterns and substantial year-to-year variation; none of the exploratory temporal trends remained statistically significant after correction for multiple comparisons. Empirical antimicrobial recommendations should therefore not be based on single-year national resistance estimates alone. Regular integration of contemporary national and local resistance surveillance into empirical treatment guidance and antimicrobial stewardship, supported by more granular and standardised susceptibility data, will be important for balancing adequate microbiological coverage with preservation of broad-spectrum antimicrobial.

## Data availability

The data analysed in this study were derived from the Iraqi national antimicrobial resistance surveillance system. The underlying surveillance data are not publicly available because they are held by the responsible national health authorities and are subject to national data governance and access restrictions. Access to the data may be considered upon reasonable request and subject to approval by the relevant Iraqi authorities and applicable data governance requirements.

## Author Contributions

**Omeed Darweesh:** Conceptualization, Methodology, Investigation, Data curation, Formal analysis, Writing–original draft, Writing–review & editing.

**Hemn Ahmed:** Investigation, Data curation, Validation, Writing – review & editing.

**Haidar Kadhim Al-Jawadi:** Investigation, Resources, Data curation, Validation, Writing – review & editing.

**Ramiar Kamal Kheder:** Methodology, Formal analysis, Validation, Writing – review & editing. **Noor Najim Alwiswasi:** Investigation, Resources, Data curation, Validation, Writing – review & editing.

**Sarah Mohammed Hameed:** Investigation, Data curation, Writing – review & editing.

**Amanj Kurdi:** Conceptualization, Methodology, Supervision, Formal analysis, Validation, Writing – original draft, Writing – review & editing, Project administration.

All authors reviewed and approved the final version of the manuscript and agree to be accountable for their contributions to the work.

**Declaration of generative AI and AI-assisted technologies in the manuscript preparation process** During the preparation of this work, the authors used ChatGPT (OpenAI) to assist with language editing, clarity, conciseness and manuscript organisation. After using this tool, the authors reviewed and edited the content as needed and take full responsibility for the content of the publication.

## List of Supplementary files

**Supplementary File 1. Resistance trajectories in *Escherichia coli*, 2021–2024**

Panels: **A, penicillins/β-lactams; B, aminoglycoside; C, macrolide; D, glycopeptide; E, tetracycline**

**Supplementary File 2. Resistance trajectories in *Klebsiella pneumoniae*, 2021–2024**

Panels: **A, penicillins/β-lactams; B, aminoglycoside; C, macrolide; D, glycopeptide; E, tetracycline**

**Supplementary File 3. Resistance trajectories in *Pseudomonas aeruginosa*, 2021–2024**

Panels: **A, aminoglycoside; B, fluoroquinolone; C, macrolide; D, glycopeptide; E, tetracycline**

Supplementary File 4. Resistance trajectories in *Staphylococcus aureus*, 2021–2024

Panels: **A, cephalosporin; B, carbapenem; C, fluoroquinolone; D, macrolide; E, tetracycline**

